# Acute Renal, Hepatic, Thromboembolic and Functional Complications after Community-Acquired Acute Lower Respiratory Tract Infection: A Prospective Cohort Study in Bristol, UK, 2022–2024

**DOI:** 10.64898/2026.08.28.26361617

**Authors:** Anastasia Chatzilena, Catherine Hyams, Robert Challen, Maria Lahuerta, Serena McGuiness, Madeleine Clout, Elizabeth Begier, Jade King, Begonia Morales-Aza, Kaltun Duale, Ainhoa Rodriguez Pereira, William Healy, Jo Southern, Philip Wells, Katie Lihou, Charli Grimes, James Campling, Nick A Maskell, Jennifer Oliver, Andrew Vyse, Bradford D. Gessner, Adam Finn, Leon Danon, the AvonCAP Research Group

## Abstract

**Introduction:** Acute lower respiratory tract disease (aLRTD) is a leading cause of hospitalisation and death, particularly in older adults and adults with comorbidities, with acute lower respiratory tract infection (aLRTI; pneumonia and non-pneumonic LRTI) being a major component. Non-pulmonary complications and functional decline after aLRTI are recognised, but their pathogen-specific burden is poorly described. We aimed to quantify renal, hepatic, thromboembolic and functional complications, and mortality, after aLRTI hospitalisation, by clinical phenotype and pathogen.

**Methods:** We conducted a cohort study of adults (≥18 years) admitted with aLRTD to two hospitals in Bristol, UK (01 August 2022–31 July 2024). aLRTD was classified as pneumonia, non-pneumonic LRTI (NP-LRTI) or no diagnosis of aLRTI. Pathogens were identified from standard-of-care and research microbiology. Outcomes were acute kidney injury (AKI), acute liver dysfunction, venous thromboembolism (VTE), in-hospital falls, reduced mobility at discharge, increased care requirements, and 30-day and 1-year mortality. Analyses were descriptive.

**Results:** Among 246,797 adult admissions, 21,456 aLRTD hospitalisations were included: 10,239 (47.7%) pneumonia, 7,742 (36.1%) NP-LRTI and 3,475 (16.2%) with no evidence of aLRTI. Of 19,152 tested aLRTD admissions, 8,503 (44.4%) had a positive microbiological/virological test, yielding 9,204 pathogen detections; 1,194 (6.2%) had co-infections, and SARS-CoV-2 was most frequent, with influenza the second most common in pneumonia and NP-LRTI. Pneumonia had greater severity than NP-LRTI and no diagnosis of aLRTI (median length of stay 6 vs 4 vs 4 days; ICU admission 3.4% vs 0.7% vs 0.5%, respectively). Overall, 22.2% developed AKI, 6.1% acute liver dysfunction, 0.6% DVT and 2.4% PE; 1.8% had a fall, 11.5% reduced mobility, and 16.6% required increased care at discharge. 30-day and 1-year mortality were highest for pneumonia (14.0% and 32.0%, respectively). Pathogen-specific analyses showed longer stays and higher complications and mortality rates for SARS-CoV-2 and *Streptococcus pneumoniae*, and shorter stays with lower complication and mortality rates for influenza and *Haemophilus influenzae*.

**Conclusions:** Non-cardiovascular complications and functional decline after aLRTI were common, particularly in pneumonic and SARS-CoV-2 or pneumococcal disease. These findings support routine surveillance for renal, hepatic, thromboembolic events, early mobilisation and rehabilitation, and consideration of multi-system outcomes when evaluating public health and economic value of vaccines and therapies.

## INTRODUCTION

Acute lower respiratory tract disease (aLRTD) remains a leading cause of adult hospitalisation and global mortality, with a particularly high burden in older adults and individuals with comorbidities (1,2). Acute lower respiratory tract infection (aLRTI) constitutes a major component of this burden. Across contemporary adult cohorts, the principal aetiologies of severe aLRTI include SARS-CoV-2, influenza viruses, respiratory syncytial virus (RSV), *Streptococcus pneumoniae* and human metapneumovirus (hMPV), all of which are frequently detected in hospitalised community-acquired pneumonia (CAP) and acute respiratory infection (ARI) (1,3–5). Vaccines targeting SARS-CoV-2, seasonal influenza, RSV, and pneumococcal disease are now recommended for many adults in national immunisation schedules, while hMPV vaccines remain in development without a licensed product (6–9). In this context, understanding the full spectrum of acute morbidity associated with these pathogens—beyond respiratory failure and short-term mortality—is increasingly important for clinical care, service planning, and assessing the public health and economic value of vaccines.

Acute non-pulmonary complications are common in adults hospitalised with pneumonia or severe aLRTI (10). Acute kidney injury (AKI) complicates approximately 18–34% of CAP admissions and independently predicts need for renal replacement therapy, prolonged length of stay and increased short- and long-term mortality (11–13). Abnormal liver function tests are likewise frequent: in CAP, hypalbuminaemia, and raised alanine aminotransferase (ALT) are associated with higher in-hospital mortality and longer stay, while in COVID-19, 40–50% of hospitalised patients exhibit liver test abnormalities that correlate with disease severity and death (14–16). Respiratory infections also confer a heightened risk of venous thromboembolism (VTE): general-practice data show a ∼2.5-fold increase in deep vein thrombosis (DVT) and pulmonary embolism (PE) in the months after respiratory infection, and recent reviews and comparative cohorts indicate that pneumonia and COVID-19 are associated with particularly high in-hospital VTE rates compared with other infections and with influenza (17–20).

In parallel, there is growing recognition that hospitalisation for influenza, pneumonia and other aLRTI can trigger sustained functional decline and increased care needs in older adults. Prospective studies show that around 18% of adults aged ≥65 years, hospitalised with influenza or other aLRTI, experience clinically meaningful loss of independence at 30 days, with nearly 10% developing catastrophic disability (21). Among older patients admitted with pneumonia, frailty strongly predicts death or functional decline within 30 days, and pneumonia hospitalisation is associated with subsequent functional and cognitive impairment, new mobility limitations and transitions to institutional care (22–24). However, most pathogen-specific work on SARS-CoV-2, influenza, and pneumococcal disease has focused on respiratory and cardiovascular endpoints, with far less attention to acute renal and hepatic dysfunction, in-hospital falls, reduced mobility, and escalation of care requirements at discharge, and very few direct comparisons of these outcomes across aetiologies. For RSV, recent studies have begun to explore non-respiratory complications in adults (25,26), but systematic cross-pathogen comparisons remain limited.

We therefore analysed a prospective cohort of adults hospitalised with community-acquired aLRTD, classifying cases as pneumonia, non-pneumonic LRTI or no clinical diagnosis of aLRTI. Within this cohort, we focused our pathogen-specific analyses on admissions with pathogen-confirmed aLRTI due to SARS-CoV-2, influenza, RSV, *S. pneumoniae*, or hMPV, and other detected pathogens to estimate the occurrence of key non-pulmonary complications—acute renal failure, acute liver dysfunction, DVT, PE, in-hospital falls, reduced mobility following admission, and increased care needs at discharge—and to compare these risks by aetiology. Our aim was to characterise the short-term multi-system burden associated with these specific respiratory pathogens and to identify patient- and pathogen-level factors associated with acute organ dysfunction, functional decline and care escalation during and immediately following aLRTD hospitalisation.

## METHODS

### Study design

We conducted a prospective observational cohort study of all adults (≥18y) hospitalised with community-acquired acute lower respiratory tract disease (aLRTD) at the two acute care hospitals in Bristol, UK between 01 August 2020 and 31 July 2024. This analysis forms part of the AvonCAP study (ISRCTN 17354061), which undertook comprehensive population-based surveillance of aLRTD within a defined geographical catchment. Full enrolment and eligibility procedures have been published previously (27) and are summarised in Supplementary Data 1 and 2. To focus on a period when respiratory virus circulation, healthcare utilisation, and testing practices had largely stabilised after the acute phases of the SARS-CoV-2 pandemic and to avoid the atypical epidemiology and admission patterns seen during earlier pandemic waves, we included in this analysis all aLRTD admitted between 01 Aug 2022 and 31 July 2024. Demographic and clinical data were collected from electronic and paper patient records and recorded using REDCap (28). Data on co-morbidities were determined at admission and included the Charlson comorbidity index (CCI) (29).

Pneumonia was classified as an acute respiratory illness with confirmed radiological changes compatible with infection or if the treating clinician made the diagnosis. In keeping with the National Institute for Health and Care Excellence (NICE) and British Thoracic Society (BTS) guidelines (30), patients assigned a pneumonia diagnosis were counted as a pneumonia case even in the absence of radiological investigation or infiltrate on imaging, due to known false-negative radiology in pneumonia. NP-LRTI was defined as aLRTI signs/symptoms without either infective radiological changes or a clinical diagnosis of pneumonia. Under these case definitions, any patients with aLRTD signs or symptoms due to non-infectious aLRTD, would have been assigned to CRDE or HF groups i.e. no diagnosis of aLRTI. Full definitions for all subgroups are provided in Supplementary Data 2.

#### Ethics and permission

The study was approved by the Health Research Authority (HRA) Research Ethics Committee (REC) East of England, Essex, REC reference 20/EE/0157. Patients with capacity provided informed consent, and declarations for participation were obtained from consultees for individuals lacking capacity. Patients who declined consent were not included in this analysis. Data were included under Section 251 of the 2006 NHS Act was used under approval from the Clinical Advisory Group (CAG) if it was not practical to approach individuals for consent.

#### Pathogen diagnosis

Standard of care microbiological testing was undertaken as part of routine clinical care by treating physicians. Testing was performed by UKHSA-accredited hospital laboratories and included respiratory virus PCR, sputum and blood culture, and urinary antigen assays. Respiratory viruses including SARS-CoV-2, influenza A/B, RSV, and hMPV were detected using multiplex PCR using either Hologic Panther® Fusion, BioFire® Diagnostics system, or Cepheid Xpert® panel tests. SARS-CoV-2 and influenza were additionally identified by Abbott® point-of-care lateral flow tests (POCT). Bacterial pathogens including *S. pneumoniae, Staphylococcus aureus,* and *Pseudomonas aeruginosa* were confirmed through standard microbiological culture or PCR of sterile-site specimens, with species identification supported by API®-20 Strep (bioMérieux) or MALDI-TOF mass spectrometry (Bruker). A positive pneumococcal urinary-antigen test [UAT] (BinaxNOW®, Alere, UK) was also considered evidence of pneumococcal infection. Patients were assigned to pathogen groups based on any positive result from these tests.

Participants who consented to the enhanced diagnostic testing study arm underwent research sampling, providing one or more of the following specimen types: an upper respiratory swab (NP or combined NP/OP swab), a saliva, or sputum sample. If subjects were unable to produce saliva, a saline mouth wash specimen was collected. Research specimens were tested by RT-qPCR using Certest Biotech Viasure® viral pathogen multiplex panels on the Applied Biosystems QuantStudio 7 Flex Real-Time PCR System (Thermo Fisher Scientific®). The sample volumes, PCR threshold and baseline values used, differed between SOC and research RT-PCR tests. Research swabs were stored in STGG (skim-milk, tryptone, glucose, glycerine) medium and SOC swabs in VTM (viral transport medium).

#### Outcomes

Pre-specified in-hospital complications were ascertained from review of medical notes, electronic laboratory results, radiology reports, drug charts and local incident-reporting systems. AKI was defined as new acute kidney injury or acute renal impairment documented by the treating clinical team during the admission according to KDIGO criteria (31). Acute liver dysfunction was defined as new-onset derangement in liver biochemistry or documented acute hepatic injury attributable to the presenting illness, identified from review of the medical records, with thresholds used in international liver injury guidance (32). Deep vein thrombosis (DVT) and pulmonary embolism (PE) were defined as radiologically confirmed venous thromboembolism events occurring during the admission (33). In-hospital falls were defined as any unintentional change in position to a lower level occurring on the ward and recorded in the medical record or incident-reporting system. Reduced mobility was defined as a new requirement for assistance with mobilising or new use of a walking aid at discharge compared with pre-admission baseline. Increased care on discharge was defined as a new or escalated formal care package, new referral to community rehabilitation services, or new discharge to a residential or nursing care facility. Further definitions are given in Supplementary Data 2.

#### Study Objectives

The primary objective was to assess the occurrence of non-cardiovascular complications within 30-days of aLRTI hospitalisation, by clinical phenotype (i.e. pneumonia, non-pneumonic aLRTI, and those without SOC evidence of aLRTI) and by pathogen. Non-cardiovascular complications included AKI, acute liver dysfunction, PE and DVT, in-hospital falls, reduced mobility, and requirement for increased care following discharge. Secondary objectives were to describe 30-day and 1-year mortality following admission and to examine basic demographic characteristics associated with these sequelae.

#### Statistical Analysis

Descriptive statistics were used to summarise in-hospital complications, discharge functional and care outcomes, post-discharge mortality (30-day and 1-year) and baseline demographics overall and stratified by both diagnosis category (pneumonia; NP-LRTI; aLRTD without SOC evidence of LRTI) and pathogen group (SARS-CoV-2; influenza; RSV; *S. pneumoniae*; human metapneumovirus; other pathogens).

Continuous variables are presented as median (interquartile range) and categorical variables as number (%). Normality of continuous data was assessed using the Anderson–Darling test. Comparisons across groups were performed using the Kruskal–Wallis test for continuous variables and Fisher’s exact test for categorical variables. Given multiple comparisons, results should be interpreted with caution; statistical significance was defined as a two-sided p-value <0.05.

Statistical analyses were conducted using R version 4.5.1 (34), and the data flow diagram was generated using the R package ‘dtrackr’(35). Missing data were reported descriptively and not imputed.

## RESULTS

Between 1 August 2022 and 31 July 2024, 246,797 adults in total were hospitalised across both acute care hospitals in Bristol, UK. Following exclusions, 21,456 unique adult aLRTD admissions were included in the analysis: 10,239 (47.7%) admissions with pneumonia; 7,742 (36.1%) NP-LRTI; and 3,475 (16.2%) admissions with no radiological or clinical evidence of aLRTI from routine clinical care (Figure 1, Supplementary Data 3).

**Figure 1.**
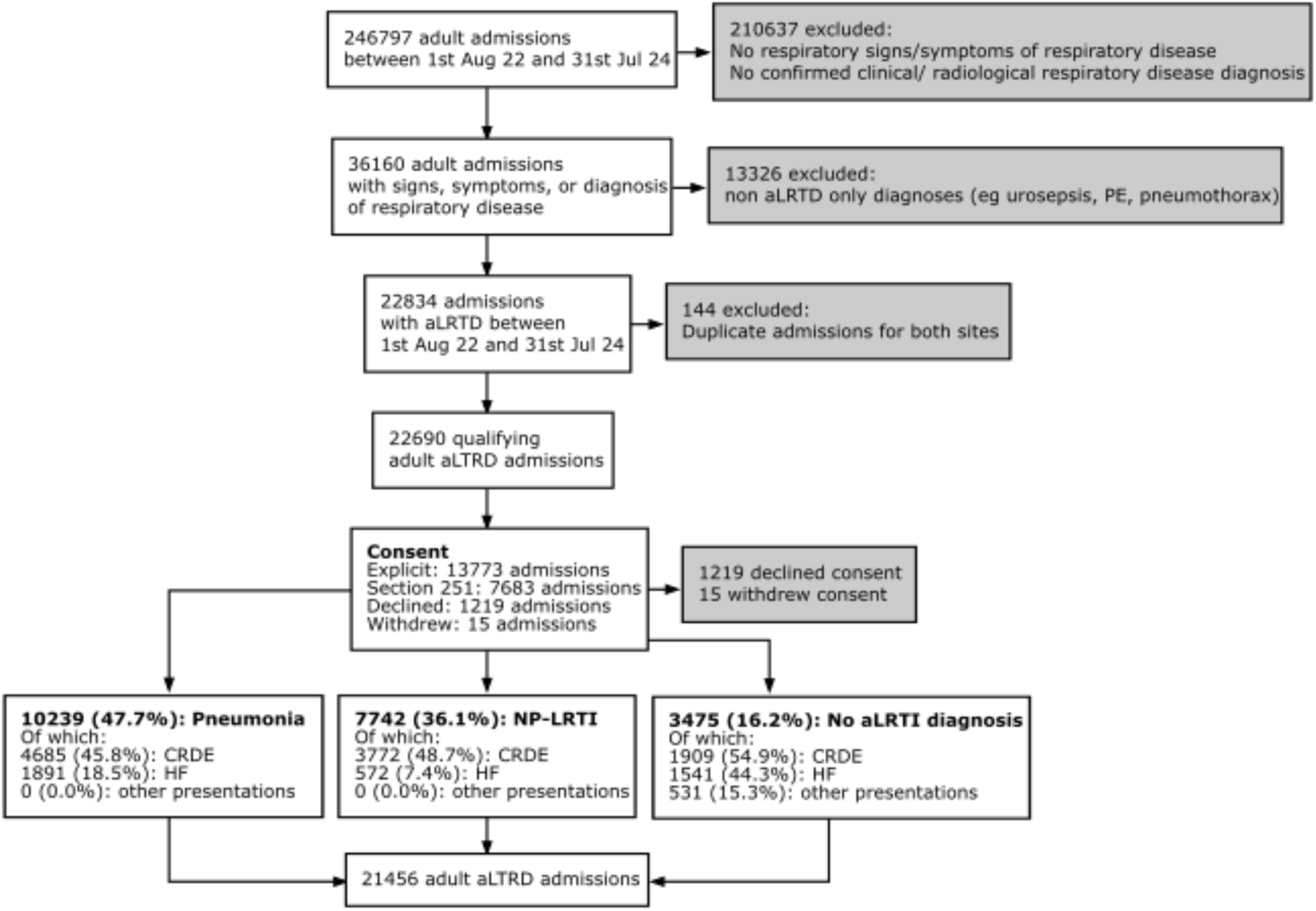
Data flow diagram. From 01 August 2022–31 July 2024, 246,797 adults were hospitalised in Bristol, UK; 36,160 had a diagnosis, signs, or symptoms of respiratory disease. After excluding non-aLRTD diagnoses (e.g. urosepsis, pulmonary embolism, pneumothorax), duplicate admissions across sites, and exclusions for declined/withdrawn consent, 21,456 unique adult aLRTD admissions were included in the analysis. These comprised 10,239 (47.7%) admissions with pneumonia, 7,742 (36.1%) with non-pneumonic lower respiratory tract infection (NP-LRTI), and 3,475 (16.2%) admissions with no diagnosis of aLRTI. aLRTD, acute lower respiratory tract disease; aLRTI, lower respiratory tract infection; CRDE, chronic respiratory disease exacerbation; HF, heart failure; NP-LRTI, non-pneumonic lower respiratory tract infection; PE, pulmonary embolus.

Of 19,152 aLRTD admissions that underwent microbiological or virological testing, 8,503 (44.4%) had at least one positive result, yielding 9,204 respiratory pathogen detections; 1,194 (6.2%) admissions had co-infections (≥2 pathogens) (Table 1, Supplementary Data 3-4). We identified 178 aLRTD cases with a culture positive for *S. epidermidis*, 227 for *E. coli*, and 363 for Candida, and we did not consider these respiratory pathogens due to the risk of contamination or infection from another source (Figure 1). The most commonly detected respiratory pathogen in all aLRTD groups was SARS-CoV-2 (Table 1) and, among cases with pneumonia and NP-LRTI, influenza was the second most common, accounting for 3.8% and 6.3% of hospitalisations, respectively. Of the 3,475 cases in which there was no suspicion of aLRTI, we found 252 cases (7.3% of cases without a clinical aLRTI diagnosis) associated with a positive microbiological test.

**Table 1.** Pathogen distribution among adult hospital aLRTD cases, stratified by pneumonia, non-pneumonic lower respiratory tract infection (NP-LRTI), and those with no aLRTI diagnosis. Values are numbers and column percentages of admissions in which each pathogen was identified within the three radiological/clinical groups. Pathogens were assigned based on standard-of-care and research microbiological testing; ‘No pathogen detected’ includes admissions with negative testing or no pathogen identified. The overlap of pathogens (i.e. instances where patients tested positive for more than one) is listed in Supplementary Data 3). The distribution of microbiological and virological test types by clinical diagnosis group is provided in Supplementary Data 4.

| Pathogen | All aLRTD<br>N=21456 | Pneumonia<br>N=10239 | NP-LRTI<br>N=7742 | No aLRTI diagnosis<br>N=3475 |
| --- | --- | --- | --- | --- |
|  | N (%) | N (%) | N (%) | N (%) |
| SARS-CoV-2 | 4444 (20.7%) | 1656 (16.2%) | 2758 (35.6%) | 30 (0.9%) |
| Influenza (A&B) | 955 (4.5%) | 426 (4.2%) | 520 (6.7%) | 9 (0.3%) |
| RSV | 384 (1.8%) | 181 (1.8%) | 202 (2.6%) | 1 (0.0%) |
| Human Metapneumovirus | 304 (1.4%) | 164 (1.6%) | 133 (1.7%) | 7 (0.2%) |
| Parainfluenza (1-4) | 269 (1.3%) | 120 (1.2%) | 142 (1.8%) | 7 (0.2%) |
| Rhinovirus/Enterovirus | 626 (2.9%) | 326 (3.2%) | 283 (3.7%) | 17 (0.5%) |
| *H. influenzae* | 420 (2.0%) | 245 (2.4%) | 141 (1.8%) | 34 (1.0%) |
| *S. pneumoniae* | 330 (1.5%) | 287 (2.8%) | 37 (0.5%) | 6 (0.2%) |
| *S. aureus* | 191 (0.9%) | 130 (1.3%) | 40 (0.5%) | 21 (0.6%) |
| *S. epidermidis* * | 178 (0.8%) | 119 (1.2%) | 45 (0.6%) | 14 (0.4%) |
| *Pseudomonas* spp | 242 (1.1%) | 152 (1.5%) | 77 (1.0%) | 13 (0.4%) |
| *Escherichia coli* * | 227 (1.1%) | 127 (1.2%) | 65 (0.8%) | 35 (1.0%) |
| *Candida* * | 363 (1.7%) | 264 (2.6%) | 73 (0.9%) | 26 (0.7%) |
| Other pathogen | 1039 (4.8%) | 686 (6.7%) | 279 (3.6%) | 74 (2.1%) |
| No pathogen detected | 10649 (49.6%) | 5575 (54.4%) | 2797 (36.1%) | 2277 (65.5%) |
| Not tested | 2304 (10.7%) | 735 (7.2%) | 623 (8.0%) | 946 (27.2%) |

\* *S. epidermidis*, *E. coli*, and *Candida* spp were not considered respiratory pathogens due to the risk of contamination or infection from another source (e.g. urinary system)
aLRTD, acute lower respiratory tract disease; aLRTI, acute lower respiratory tract infection; hMPV, human metapneumovirus; NP-LRTI, non-pneumonic lower respiratory tract infection; RSV, respiratory syncytial virus

The demographics of patients hospitalised with aLRTD by clinical phenotype are presented in Table 2. Patients hospitalised with pneumonia had longer admissions (6 days [IQR:2-14] vs 4 days [IQR:1-10] for NP-LRTI and 4 days [IQR:1-9] in those with no diagnosis of aLRTI) (Table 3). Markers of acute severity were consistently highest in patients with pneumonia compared with those with NP-LRTI and those without a diagnosis of aLRTI: ICU admission (3.4% vs 0.7% and 0.5%), need for any respiratory support (31.6% vs 19.9% and 21.5%), and use of advanced ventilatory modalities, including intubation (1.7% vs 0.2% and 0.2%). Pneumonia was also associated with the greatest burden of acute renal failure, liver dysfunction, reduced mobility, and increased care needs at discharge. Short- and longer-term outcomes were poorest for pneumonia, with 30-day and 1-year mortality of 14.0% and 32.0%, compared with 3.8% and 16.5% for NP-LRTI and 6.7% and 22.6%, respectively, for patients with no diagnosis of aLRTI.

**Table 2.**
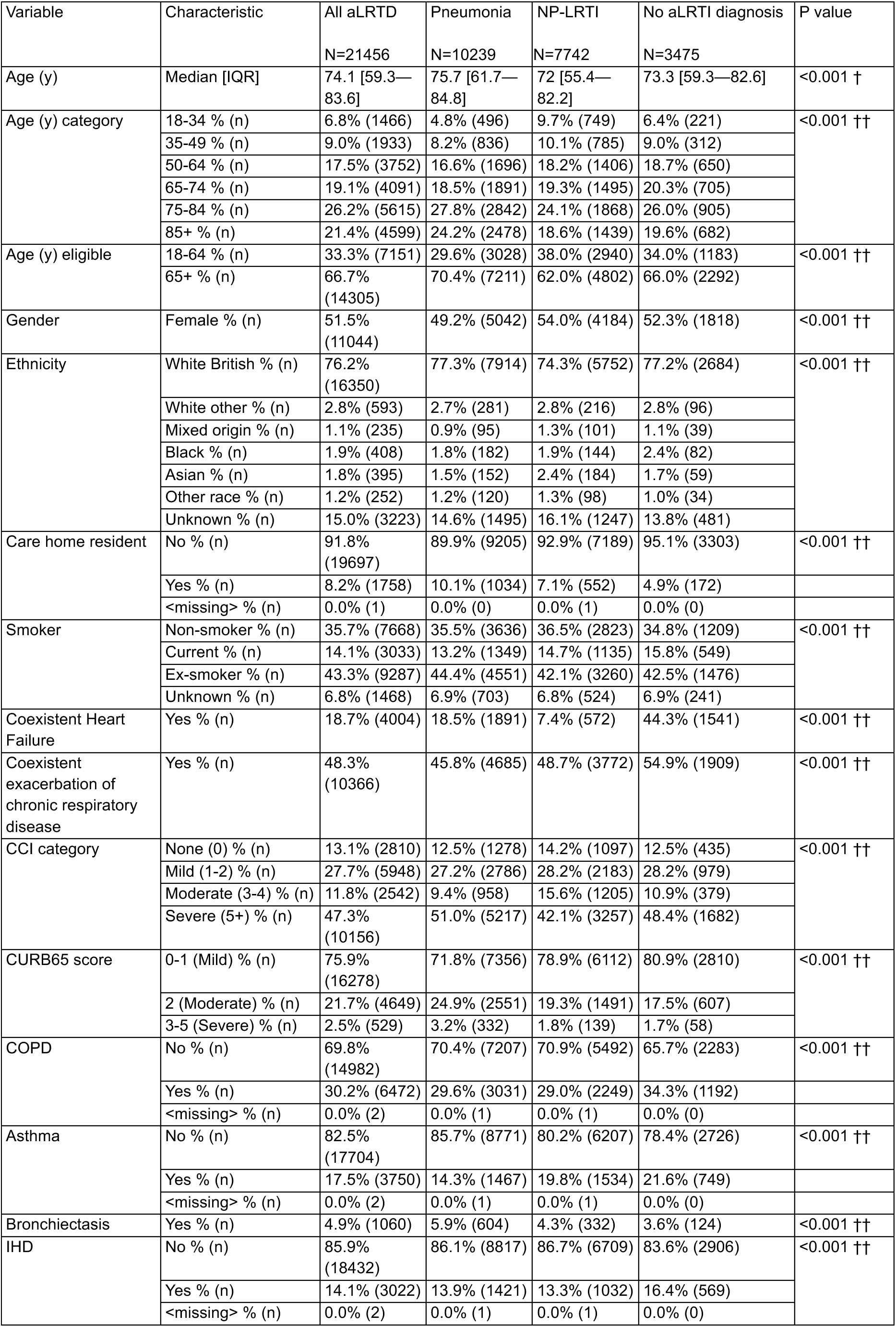
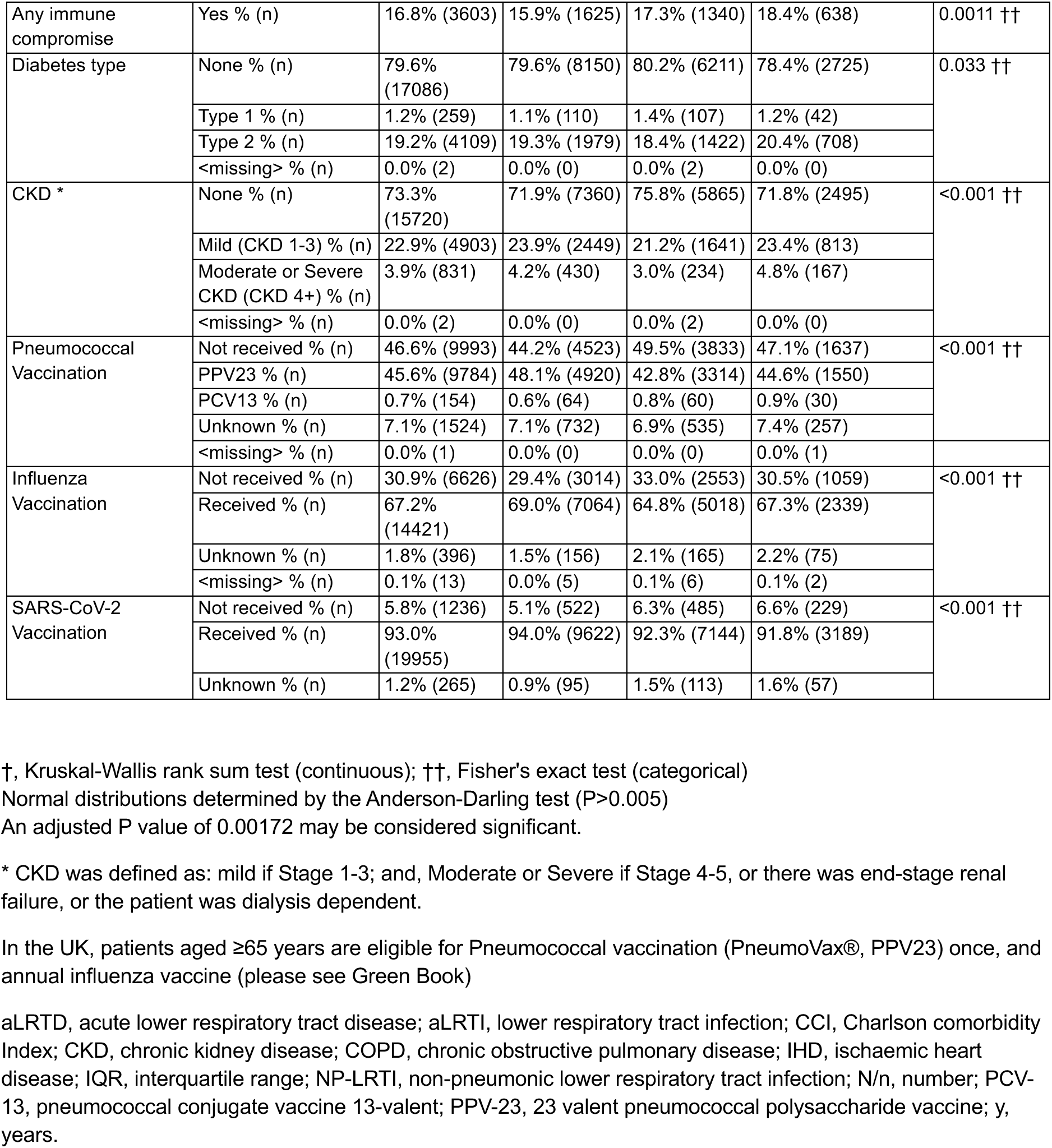
Demographic characteristics of patients hospitalised with aLRTD, stratified by clinical diagnosis.

**Table 3.** Outcomes in the 30-days after hospital admission for patients hospitalised with aLRTD, stratified by clinical diagnosis.

| Variable | Characteristic | All aLRTD<br>N=21456 | Pneumonia<br>N=10239 | NP-LRTI<br>N=7742 | No aLRTI<br>diagnosis<br>N=3475 | P value |
| --- | --- | --- | --- | --- | --- | --- |
| Length of stay | Median [IQR] | 5 [2—11] | 6 [2—14] | 4 [1—10] | 4 [1—9] | <0.001 † |
| ICU admission | Yes % (n) | 2.0% (426) | 3.4% (350) | 0.7% (57) | 0.5% (19) | <0.001 †† |
|  | No % (n) | 98.0% (21030) | 96.6% (9889) | 99.3% (7685) | 99.5% (3456) |  |
| ICU admission duration | Median [IQR] | 6 [3—11] | 7 [3.5—12] | 3 [2—7] | 4 [2—7] | — ††† |
| Respiratory support needed | No % (n) | 74.2% (15921) | 68.3% (6994) | 80.1% (6199) | 78.5% (2728) | <0.001 †† |
|  | Yes % (n) | 25.7% (5518) | 31.6% (3235) | 19.9% (1537) | 21.5% (746) |  |
|  | <missing> % (n) | 0.1% (17) | 0.1% (10) | 0.1% (6) | 0.0% (1) |  |
| Highest level ventilatory support | Intubation % (n) | 0.9% (195) | 1.7% (175) | 0.2% (14) | 0.2% (6) | <0.001 †† |
|  | BiPAP % (n) | 3.8% (818) | 4.5% (463) | 2.9% (228) | 3.7% (127) |  |
|  | CPAP % (n) | 2.9% (624) | 3.5% (355) | 2.2% (171) | 2.8% (98) |  |
|  | High-Flow Nasal Cannulae % (n) | 6.9% (1482) | 8.7% (890) | 5.6% (434) | 4.5% (158) |  |
|  | None % (n) | 85.5% (18337) | 81.6% (8356) | 89.1% (6895) | 88.8% (3086) |  |
| Ventilator duration | Median [IQR] | 4 [2—6] | 4 [2—7] | 3 [2—5] | 3 [2—5] | — ††† |
| Renal support duration | Median [IQR] | 0 [0—0] | 0 [0—0] | 0 [0—0] | 0 [0—0] | — ††† |
| Acute renal failure | Yes % (n) | 22.2% (4754) | 26.4% (2704) | 17.7% (1370) | 19.6% (680) | <0.001 †† |
| Liver dysfunction | Yes % (n) | 6.1% (1308) | 7.4% (762) | 4.8% (371) | 5.0% (175) | <0.001 †† |
| DVT | Yes % (n) | 0.6% (119) | 0.6% (64) | 0.4% (29) | 0.7% (26) | 0.016 †† |
| PE | Yes % (n) | 2.4% (523) | 2.4% (250) | 1.0% (80) | 5.6% (193) | <0.001 †† |
| Fall in hospital | Yes % (n) | 1.8% (396) | 1.9% (197) | 2.1% (165) | 1.0% (34) | <0.001 †† |
| Reduced mobility | Yes % (n) | 11.5% (2467) | 13.2% (1352) | 10.4% (807) | 8.9% (308) | <0.001 †† |
| Increasing care requirement | Yes % (n) | 16.6% (3563) | 17.5% (1796) | 16.4% (1268) | 14.4% (499) | <0.001 †† |
| Functional status | Deceased % (n) | 9.4% (2017) | 14.3% (1467) | 4.0% (313) | 6.8% (237) | <0.001 †† |
|  | Recovered % (n) | 77.1% (16534) | 70.5% (7218) | 83.7% (6483) | 81.5% (2833) |  |
|  | Recovered with long term sequelae % (n) | 0.8% (178) | 0.8% (84) | 0.8% (59) | 1.0% (35) |  |
|  | Ongoing recovery % (n) | 4.1% (878) | 4.6% (469) | 3.4% (266) | 4.1% (143) |  |
|  | Not recovered % (n) | 8.5% (1834) | 9.7% (996) | 8.0% (616) | 6.4% (222) |  |
|  | Unknown % (n) | 0.1% (14) | 0.0% (5) | 0.1% (4) | 0.1% (5) |  |
|  | <missing> % (n) | 0.0% (1) | 0.0% (0) | 0.0% (1) | 0.0% (0) |  |
| Death within 30 days | Yes % (n) | 9.1% (1954) | 14.0% (1429) | 3.8% (291) | 6.7% (234) | <0.001 †† |
|  | No % (n) | 90.9% (19502) | 86.0% (8810) | 96.2% (7451) | 93.3% (3241) |  |
| Death within 1 year | Yes % (n) | 24.9% (5338) | 32.0% (3274) | 16.5% (1280) | 22.6% (784) | <0.001 †† |
|  | No % (n) | 75.1% (16118) | 68.0% (6965) | 83.5% (6462) | 77.4% (2691) |  |

†, Kruskal-Wallis rank sum test (continuous); ††, Fisher's exact test (categorical); †††, Not calculated due to missing values (continuous)
Normal distributions determined by the Anderson-Darling test ( $P>0.005$ )
aLRTD, acute lower respiratory tract disease; aLRTI, lower respiratory tract infection; BiPAP, Bilevel Positive Airway Pressure; CPAP, Continuous Positive Airways Pressure; ICU, intensive care unit; IQR, interquartile range; NP-LRTI, non-pneumonic lower respiratory tract infection; N/n, number

The demographics of patients hospitalised with aLRTD by pathogen are presented in Table 4. Across pathogen groups, severity and outcomes differed (Table 5). Median length of stay differed across pathogen groups (p<0.001), being longest for SARS-CoV-2, pneumococcal and “other detected pathogen” infections (7–9 days) and shorter for influenza, *H. influenzae*, rhinovirus/enterovirus, RSV, hMPV and parainfluenza (4–5 days). The “other detected pathogen” group comprised admissions with Pseudomonas spp., *Staphylococcus aureus*, or pathogens classified as ‘Other pathogen’ in Table 1 (pathogens detected on testing and were not included in the named strata). ICU admission and advanced respiratory support were also concentrated in pneumococcal and ‘other detected pathogen’ cases, with intermediate use in hMPV and SARS-CoV-2 and lowest rates in influenza and *H. influenzae* (p<0.001). Acute renal failure and liver dysfunction were most frequent in SARS-CoV-2, pneumococcal and ‘other detected pathogen’ infections, and less common in influenza and *H. influenzae* (p≤0.015). Functional sequelae were greatest for SARS-CoV-2 and “other pathogen” groups, which had the highest rates of in-hospital falls, reduced mobility and increased care requirements at discharge. We found 30-day and 1-year mortality were highest in ‘other detected pathogen’ and SARS-CoV-2 infections and lowest after influenza and *H. influenzae* (p<0.001).

**Table 4.**
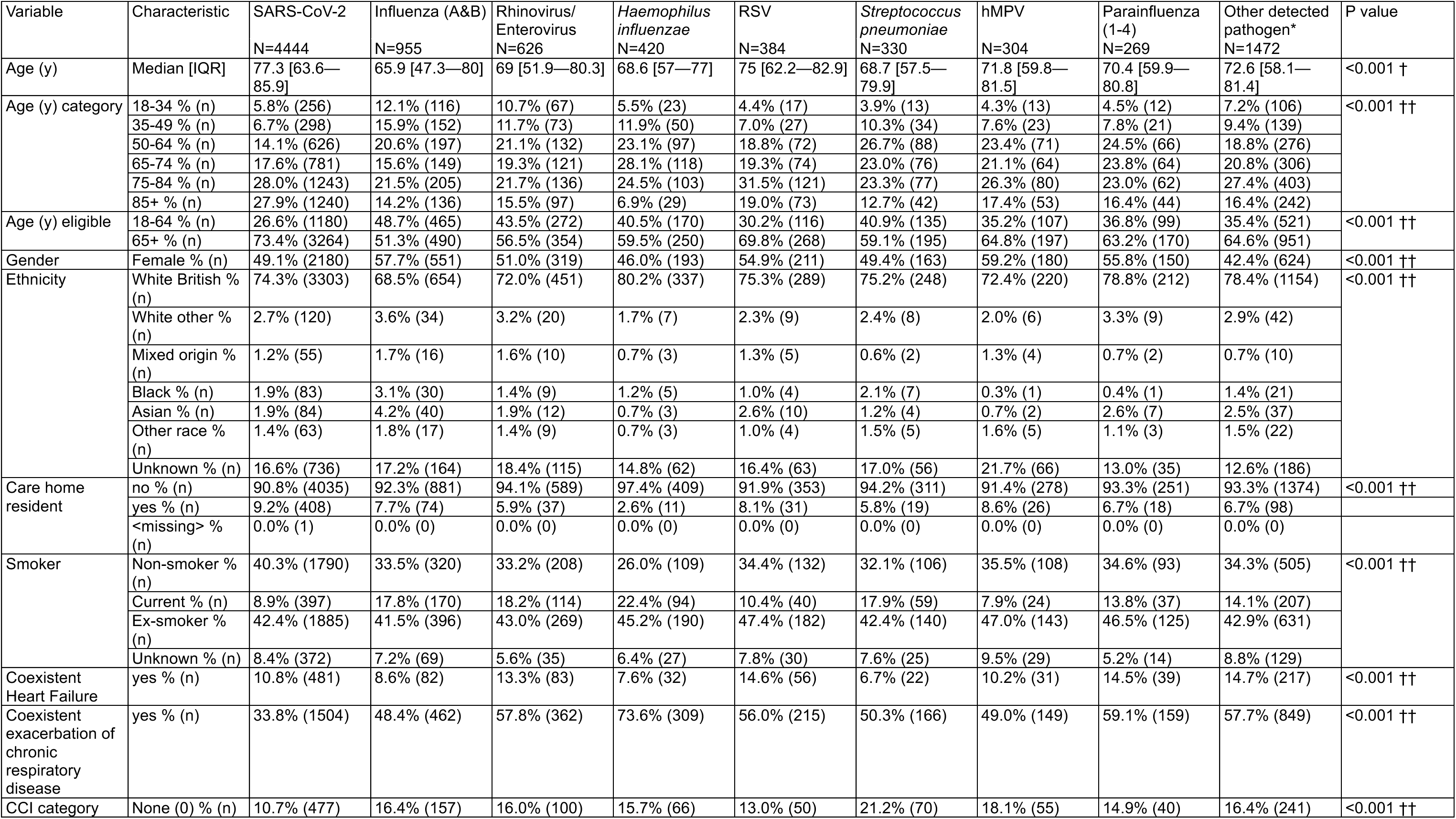
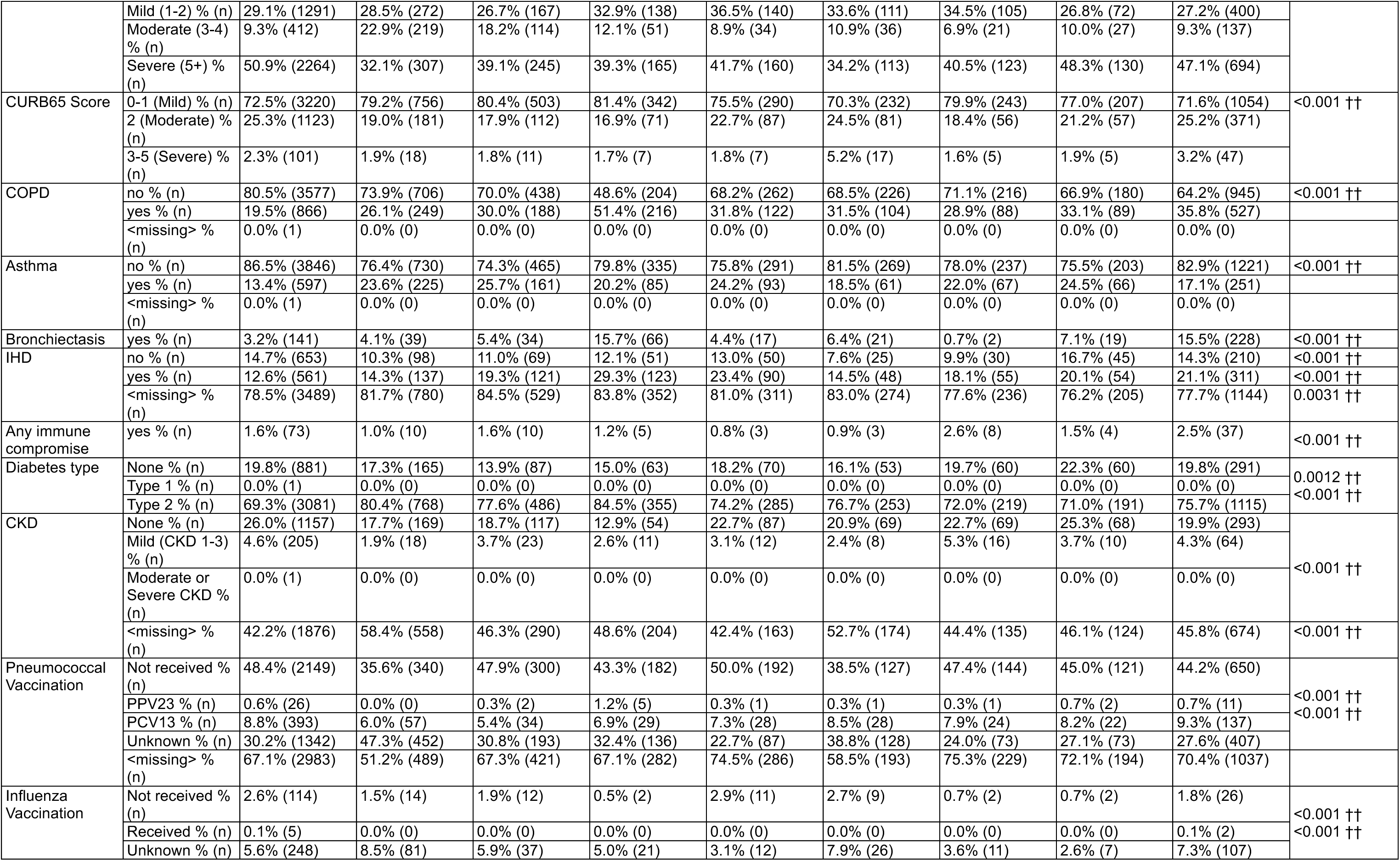
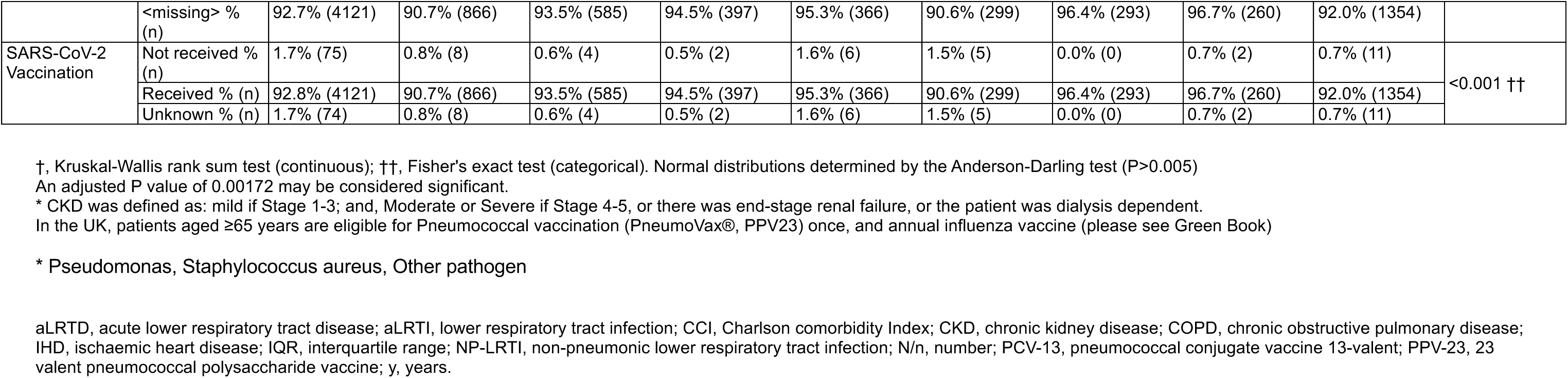
Demographic characteristics of patients hospitalised with aLRTD and positive microbiology and virology tests, stratified by pathogen.

**Table 5.** Outcomes in the 30-days after hospital admission for patients hospitalised with aLRTD and positive microbiology and virology tests, stratified by pathogen.

| Variable | Characteristic | SARS-CoV-2<br>N=4444 | Influenza (A&B)<br>N=955 | Rhinovirus/<br>Enterovirus<br>N=626 | Haemophilus influenzae<br>N=420 | RSV<br>N=384 | Streptococcus pneumoniae<br>N=330 | hMPV<br>N=304 | Parainfluenza (1-4)<br>N=269 | Other detected pathogen*<br>N=1472 | P value |
| --- | --- | --- | --- | --- | --- | --- | --- | --- | --- | --- | --- |
| Length of stay | Median [IQR] | 9 [2—23] | 4 [1—8] | 5 [2—10] | 4 [2—9] | 5 [2—10] | 7 [4—14] | 5 [2—9] | 5 [2—11] | 9 [4—20] | <0.001 † |
| ICU admission | Yes % (n) | 2.8% (123) | 2.7% (26) | 3.5% (22) | 3.1% (13) | 2.9% (11) | 11.2% (37) | 4.9% (15) | 3.0% (8) | 9.6% (142) | <0.001 †† |
|  | No % (n) | 97.2% (4321) | 97.3% (929) | 96.5% (604) | 96.9% (407) | 97.1% (373) | 88.8% (293) | 95.1% (289) | 97.0% (261) | 90.4% (1330) |  |
| ICU admission duration | Median [IQR] | 6 [3—10.5] | 8 [4.25—12.8] | 5.5 [3—9.5] | 7 [5—13] | 6 [2.5—21.5] | 6 [3—12] | 4 [2—10.5] | 16.5 [4.75—31.5] | 10 [5—18] | — ††† |
| Respiratory support needed | No % (n) | 79.8% (3546) | 71.9% (687) | 70.4% (441) | 67.1% (282) | 66.9% (257) | 60.6% (200) | 67.4% (205) | 68.0% (183) | 60.9% (896) | <0.001 †† |
|  | Yes % (n) | 20.1% (892) | 28.1% (268) | 29.6% (185) | 32.9% (138) | 33.1% (127) | 39.4% (130) | 32.6% (99) | 32.0% (86) | 39.1% (575) |  |
|  | <missing> % (n) | 0.1% (6) | 0.0% (0) | 0.0% (0) | 0.0% (0) | 0.0% (0) | 0.0% (0) | 0.0% (0) | 0.0% (0) | 0.1% (1) |  |
| Highest level ventilatory support | Intubation % (n) | 1.3% (57) | 1.4% (13) | 1.1% (7) | 2.1% (9) | 1.3% (5) | 5.5% (18) | 1.6% (5) | 2.2% (6) | 5.6% (82) | <0.001 †† |
|  | BiPAP % (n) | 2.6% (117) | 3.0% (29) | 4.6% (29) | 3.1% (13) | 3.9% (15) | 3.3% (11) | 4.9% (15) | 3.7% (10) | 7.4% (109) |  |
|  | CPAP % (n) | 3.0% (132) | 2.4% (23) | 3.5% (22) | 2.9% (12) | 4.2% (16) | 3.6% (12) | 3.3% (10) | 4.1% (11) | 5.0% (73) |  |
|  | High-Flow Nasal Cannulae % (n) | 5.8% (258) | 6.4% (61) | 8.3% (52) | 12.4% (52) | 10.4% (40) | 11.5% (38) | 11.5% (35) | 8.2% (22) | 9.9% (145) |  |
|  | None % (n) | 87.3% (3880) | 86.8% (829) | 82.4% (516) | 79.5% (334) | 80.2% (308) | 76.1% (251) | 78.6% (239) | 81.8% (220) | 72.2% (1063) |  |
| Ventilator duration | Median [IQR] | 5 [2—8] | 4 [2—6] | 4 [2—6] | 3 [1—5] | 3 [2—5] | 4 [2—7] | 3 [2—5] | 4 [2—6] | 5 [3—10] | — ††† |
| Renal support duration | Median [IQR] | 0 [0—0] | 0 [0—0] | 0 [0—0] | 0 [0—0] | 0 [0—0] | 0 [0—0] | 0 [0—0] | 0 [0—0] | 0 [0—0] | — ††† |
| Acute renal failure | Yes % (n) | 27.4% (1218) | 19.2% (183) | 14.7% (92) | 11.7% (49) | 25.3% (97) | 30.0% (99) | 22.4% (68) | 20.4% (55) | 29.0% (427) | <0.001 †† |
| Liver dysfunction | Yes % (n) | 6.5% (291) | 7.1% (68) | 5.8% (36) | 4.5% (19) | 6.2% (24) | 9.4% (31) | 7.9% (24) | 3.7% (10) | 8.4% (123) | 0.015 †† |
| DVT | Yes % (n) | 0.6% (28) | 0.0% (0) | 0.5% (3) | 1.2% (5) | 0.0% (0) | 0.6% (2) | 0.3% (1) | 0.4% (1) | 1.0% (15) | 0.018 †† |
| PE | Yes % (n) | 1.6% (71) | 0.7% (7) | 1.0% (6) | 2.1% (9) | 1.0% (4) | 1.5% (5) | 1.3% (4) | 0.4% (1) | 2.8% (41) | 0.0042 †† |
| Fall in hospital | Yes % (n) | 3.9% (173) | 1.5% (14) | 0.8% (5) | 0.7% (3) | 1.8% (7) | 0.6% (2) | 1.0% (3) | 1.5% (4) | 2.0% (30) | <0.001 †† |
| Reduced mobility | Yes % (n) | 18.2% (810) | 8.3% (79) | 7.0% (44) | 5.0% (21) | 8.1% (31) | 8.5% (28) | 6.2% (19) | 8.6% (23) | 13.9% (204) | <0.001 †† |
| Increasing care requirement | Yes % (n) | 24.3% (1078) | 11.3% (108) | 15.0% (94) | 12.4% (52) | 12.8% (49) | 14.5% (48) | 11.2% (34) | 13.8% (37) | 19.7% (290) | <0.001 †† |
| Functional status | Deceased % (n) | 8.7% (388) | 4.9% (47) | 4.8% (30) | 3.8% (16) | 6.0% (23) | 7.6% (25) | 6.9% (21) | 5.2% (14) | 12.2% (180) | <0.001 †† |
|  | Recovered % (n) | 66.8% (2967) | 86.6% (827) | 84.7% (530) | 86.7% (364) | 85.7% (329) | 79.7% (263) | 84.9% (258) | 82.5% (222) | 63.8% (939) |  |
|  | Recovered with long term sequelae % (n) | 1.0% (46) | 0.6% (6) | 0.6% (4) | 1.2% (5) | 0.8% (3) | 0.9% (3) | 0.0% (0) | 0.4% (1) | 1.3% (19) |  |
|  | Ongoing recovery % (n) | 4.9% (216) | 2.9% (28) | 2.9% (18) | 4.8% (20) | 2.9% (11) | 5.5% (18) | 2.3% (7) | 1.9% (5) | 6.0% (88) |  |
|  | Not recovered % (n) | 18.6% (825) | 4.9% (47) | 7.0% (44) | 3.6% (15) | 4.7% (18) | 6.1% (20) | 5.9% (18) | 10.0% (27) | 16.6% (245) |  |
|  | Unknown % (n) | 0.0% (2) | 0.0% (0) | 0.0% (0) | 0.0% (0) | 0.0% (0) | 0.3% (1) | 0.0% (0) | 0.0% (0) | 0.1% (1) |  |
| Death within 30 days | Yes % (n) | 7.7% (343) | 4.9% (47) | 4.8% (30) | 3.6% (15) | 6.0% (23) | 7.6% (25) | 6.9% (21) | 4.8% (13) | 11.7% (172) | <0.001 †† |
|  | No % (n) | 92.3% (4101) | 95.1% (908) | 95.2% (596) | 96.4% (405) | 94.0% (361) | 92.4% (305) | 93.1% (283) | 95.2% (256) | 88.3% (1300) |  |
| Death within 1 year | Yes % (n) | 26.3% (1168) | 12.4% (118) | 15.5% (97) | 11.9% (50) | 16.1% (62) | 18.2% (60) | 14.1% (43) | 16.7% (45) | 31.1% (458) | <0.001 †† |
|  | No % (n) | 73.7% (3276) | 87.6% (837) | 84.5% (529) | 88.1% (370) | 83.9% (322) | 81.8% (270) | 85.9% (261) | 83.3% (224) | 68.9% (1014) |  |

†, Kruskal-Wallis rank sum test (continuous); ††, Fisher's exact test (categorical); †††, Not calculated due to missing values (continuous)
Normal distributions determined by the Anderson-Darling test (P>0.005)
\* Pseudomonas, Staphylococcus aureus, Other pathogen
aLRTD, acute lower respiratory tract disease; aLRTI, lower respiratory tract infection; BiPAP, Bilevel Positive Airway Pressure; CPAP, Continuous Positive Airways Pressure; ICU, intensive care unit; IQR, interquartile range; NP-LRTI, non-pneumonic lower respiratory tract infection; N/n, number

## DISCUSSION

Our findings highlight a substantial multi-system burden associated with adult aLRTI that extends beyond respiratory failure and short-term death. Across more than 21,000 admissions, around 20% of patients developed AKI and >10% experienced reduced mobility or required increased care at discharge, with consistently worse outcomes in clinically diagnosed pneumonia compared with NP-LRTI and those without routine evidence of aLRTI. These data confirm that aLRTI is not only an acute pulmonary illness but a systemic insult with important renal, hepatic, thromboembolic and functional sequelae, and they show that this burden varies both by clinical phenotype and by pathogen.

The observed AKI frequency in our cohort is in keeping with previous pneumonia studies. We found AKI in 22.2% of all aLRTD admissions and in 27–30% of patients with SARS-CoV-2, pneumococcal or ‘other’ pathogen infection. Prior work in CAP has reported AKI incidences of approximately 18–34%, and has shown that even modest creatinine rises independently predict need for mechanical ventilation, renal replacement therapy, and increased short- and long-term mortality (11,36,37). Our findings therefore reinforce the concept of a lung–kidney axis in acute infection, likely mediated by systemic inflammation, haemodynamic instability, nephrotoxic exposures and microvascular injury, and they extend this evidence across a broad spectrum of viral and bacterial aetiologies (38).

Acute liver dysfunction was less frequent than AKI but still occurred in around 6% of admissions, with higher rates in pneumonia and in SARS-CoV-2, pneumococcal and ‘other’ infections. This aligns with older CAP work showing that hypoalbuminaemia and raised ALT are common and associated with higher in-hospital mortality and prolonged length of stay (14). In COVID-19, meta-analyses and cohort studies suggest that 25–40% of hospitalised patients have abnormal liver tests, and that liver biochemistry derangement is linked to greater disease severity and death (15,16,39). Our pathogen-stratified results are consistent with these observations and indicate that clinically recognised hepatic involvement is not confined to SARS-CoV-2, but also characterises pneumococcal and mixed/other infections. This supports routine monitoring of liver tests during aLRTI admission and suggests that combined renal and hepatic dysfunction may be a simple bedside marker of systemic injury.

Functional and care outcomes were strikingly common. Across all aLRTD phenotypes, 11.5% of patients had reduced mobility at discharge and 16.6% required increased formal care, with highest rates in pneumonia and in SARS-CoV-2 or ‘other pathogen’ infections. These findings are closely aligned with prospective data in older adults hospitalised with influenza or other aLRTI, where approximately 18% experience clinically important functional decline at 30 days and around 10% develop catastrophic disability (21) and echoing studies showing that RSV and pneumonia admission is associated with new impairments in activities of daily living, longer-term sequelae and increased care needs after discharge, and, for pneumonia specifically, with cognitive decline, depression and subsequent institutionalisation (23,25,26,40). By capturing in-hospital falls, new mobility limitations and escalation of care packages, our study reinforces that functional trajectories and social care needs are central outcomes of aLRTI, not merely secondary considerations.

The clear gradient in severity between pneumonia, NP-LRTI and patients without clinical evidence of aLRTI is clinically intuitive yet important to quantify. Pneumonia admissions in our cohort had longer length of stay, greater ICU use and consistently higher rates of organ dysfunction and functional decline, with 30-day and 1-year mortality of 14.0% and 32.0% respectively. These long-term mortality figures are very similar to previous CAP cohorts reporting one-year mortality approaching or exceeding 30% despite apparent resolution of the acute episode (41,42). The NP-LRTI and clinically suspected but unconfirmed aLRTI also carried substantial one-year mortality (16.5% and 22.6%) underscoring that even ‘milder’ aLRTI presentations herald a period of increased vulnerability and should trigger systematic follow-up, particularly in older and comorbid patients.

Pathogen-specific analyses showed that *S. pneumoniae* and ‘other detected pathogen’ infections had the highest ICU admission and respiratory support rates, while SARS-CoV-2, *S. pneumoniae* and ‘other detected pathogen’ were associated with longer stays, higher rates of AKI and hepatotoxicity, and the poorest functional and mortality outcomes. RSV and hMPV generally showed intermediate severity and complication profiles, not reaching the high levels seen with SARS-CoV-2, *S. pneumoniae* and ‘other detected pathogens’. Of note, this study spanned pandemic and post-pandemic periods, with substantial temporal variation in public health measures, circulating SARS-CoV-2 variants and vaccine coverage. In contrast, influenza and *H. influenzae* had shorter stays, fewer complications, and lower 30-day and 1-year mortality. These patterns are broadly consistent with comparative studies reporting that, COVID-19 and RSV or pneumococcal disease often cause more severe acute illness than influenza in hospitalised adults (26,43–45). The high burden of complications in pneumococcal aLRTI is biologically plausible given experimental evidence that *S. pneumoniae* can invade the myocardium and other organs, forming microlesions that disrupt tissue function (46). Our data extend this by demonstrating a similarly disproportionate renal and functional burden, supporting a view of pneumococcal respiratory infection as a systemic disease rather than a purely pulmonary one.

These results have several strengths, and these have been previously published (27,47). The study used population-based surveillance across a defined catchment with prospective enrolment and systematic capture of clinical, microbiological and outcome data, including enhanced molecular diagnostics for a broad pathogen panel. Pre-specified, clinically grounded definitions of AKI, liver dysfunction, VTE, falls, mobility and care escalation were applied consistently, and outcomes were examined both by clinical phenotype and by pathogen. However, limitations must be acknowledged. As an observational study, causal inference is not possible and residual confounding by frailty, baseline function or unmeasured comorbidities is likely. Pathogen ascertainment depended partly on clinician-instigated testing, which may have varied over time and by severity, leading to misclassification or under-detection (Supplementary Data 4). Some pathogens, particularly hMPV and parainfluenza, were relatively infrequent, so estimates for these groups are imprecise. This study used a combination of standard-of-care and research testing and the frequency and sensitivity of testing for various pathogens differed on this basis. On this basis, the relative frequency of specific pathogen cases would not be representative of true pathogen frequency. Finally, we captured complications during the index admission to 30-days, and vital status to one year, but did not collect more detailed post-discharge trajectories of renal recovery, liver function or functional status.

From a clinical and policy perspective, our findings argue for a broader framing of aLRTI severity that explicitly includes acute renal and hepatic injury, VTE, falls, functional decline and care escalation. These outcomes are highly relevant to patients and health systems, and appear to differ by pathogen, which has implications for vaccine policy and for the design of future trials. For example, as adult RSV and next-generation pneumococcal and COVID-19 vaccines are deployed, prevention of AKI, functional loss and transitions to higher levels of care may prove as important as reductions in respiratory failure or short-term mortality (45,48) and should therefore be included in vaccine impact models and cost-benefit calculations which underpin policy decisions. Future work should therefore prioritise longitudinal assessment of organ function and disability after aLRTI, incorporate frailty and baseline functional measures into risk stratification, and evaluate pathogen-specific prevention and rehabilitation strategies that address the full multi-system burden of acute respiratory infection in adults.

## CONCLUSIONS

Hospitalisation with aLRTI is associated with a substantial burden that extends beyond respiratory failure and short-term mortality. Acute renal and hepatic dysfunction, thromboembolic complications, functional decline and increased care needs were common, particularly among patients with pneumonia, and varied by infecting pathogen. These findings support assessment and prevention strategies that address the broader consequences of acute respiratory infection in adults.

## Data Availability

The data used in this study are sensitive and cannot be made publicly available without breaching patient confidentiality rules. Therefore, individual participant data and a data dictionary are not available to other researchers.

## ACKNOWLEDGEMENTS

We thank colleagues for their support with this study, including Rachel Davies, Paul Savage, Emma Foose, Susan Christie, Mark Mummé, and Adam Taylor. We also thank the research teams at North Bristol and University Hospitals of Bristol and Weston NHS Trusts for making this study possible, including Helen Lewis-White, Rebecca Smith, Rajeka Lazarus, Mark Lyttle, Kelly Turner, Jane Blazeby, Diana Benton, and David Wynick.

## DATA SHARING

The data from this study are sensitive and cannot be made publicly available without breaching patient confidentiality rules. The data dictionary is therefore unavailable.

## CONTRIBUTORS

CH, EB, JS, BG and AF designed the study. AC, CH, ML, CT, EB, BG, LD, and AF generated the research questions and analysis plan. CH, JK, SM, DA, and The AvonCAP team were involved in data collection. AC, CH, RC, PW, KL, LD, and AF undertook data analysis. All authors were involved in the final manuscript preparation and its revisions before publication. The data were verified by CH, DA, SM, and JK. AF provided oversight of the research. CH and AF act as guarantors of the research data.

## THE AvonCAP RESEARCH GROUP

Aaran Sinclair, Ainhoa Rodriguez-Pereira, Amelia Langdon, Amy Taylor, Anabella Turner, Anna Jones, Anna Koi, Anya Mattocks, Begonia Morales-Aza, Bethany Osborne, Brianna Dooley, Callum Hawkins, Charli Grimes, Chloe Farren, Christian Povey, Claire Mitchell, David Adegbite, Dylan Thomas, Elinor Balch, Ella Ackroyd-Weldon, Emma Bridgeman, Emma Scott, Felix Wright, Ffion Davies, Fiona Perkins, Francesca Bayley, Gabriella Ruffino, Gabriella Valentine, Georgina Mortimer, Grace Tilzey, Harriet Ibbotson, Hanah Batholomew, Hugo Swift, Jacob Symanowski, Ilana Kelland, Imogen Ely, Jade King, Jake Whittle, Jane Kinney, Johanna Kellett Wright, Jonathan Vowles, Josephine Bonnici, Josh Anderson, Juan Garcia-Tello, Julia Brzezinska, Julie Cloake, Kaltun Duale, Katarina Milutinovic, Kate Helliker, Katie Maughan, Kazminder Fox, Kellie Pettinger, Konstantina Minou, Kyla Chandler, Lana Ward, Leah Fleming, Leigh Morrison, Liberty Smith, Lily Smart, Lisa Grimmer, Louise Setter, Louise Wright, Lucy Grimwood, Maddalena Bellavia, Madeleine Clout, Maia Lyall, Malak Eghleilib, Marianne Vasquez, Maria Garcia Gonzalez, Mariella Ardeshir, Marta Mergulhao, Martina Chmelarova, Matthew Randell, Michael Booth, Milo Jeenes-Flanagan, Miriama Resutikova, Monika Chaulagain, Natalie Chang, Nefeli Tavira, Nellie Farhoudi, Niall Grace, Nicola Manning, Oliver Griffiths, Olivia Pearce, Pip Croxford, Peter Sequenza, Petronela Anchidin, Rajeka Lazarus, Rebecca Clemence, Rosa Aldridge, Rhian Walters, Riley Cooper, Robin Marlow, Robyn Heath, Rupert Antico, Sandi Nammuni Arachchge, Sarah Stollery, Seevakumar Suppiah, Sean Robinson, Serena McGuinness, Siddiqa Uddin, Taslima Mona, Tawassal Riaz, Teagan Barrett, Tom Long, Tudor Dimofte, William Healy, Yassin Ben Khoud, Vicki Mackay, Zahra Hashmi, Zandile Maseko, Zoe Taylor, Zsuzsa Szasz-Benczur, Zsolt Friedrich.

## SUPPLEMENTARY DATA

**SUPPLEMENTARY DATA 1:**
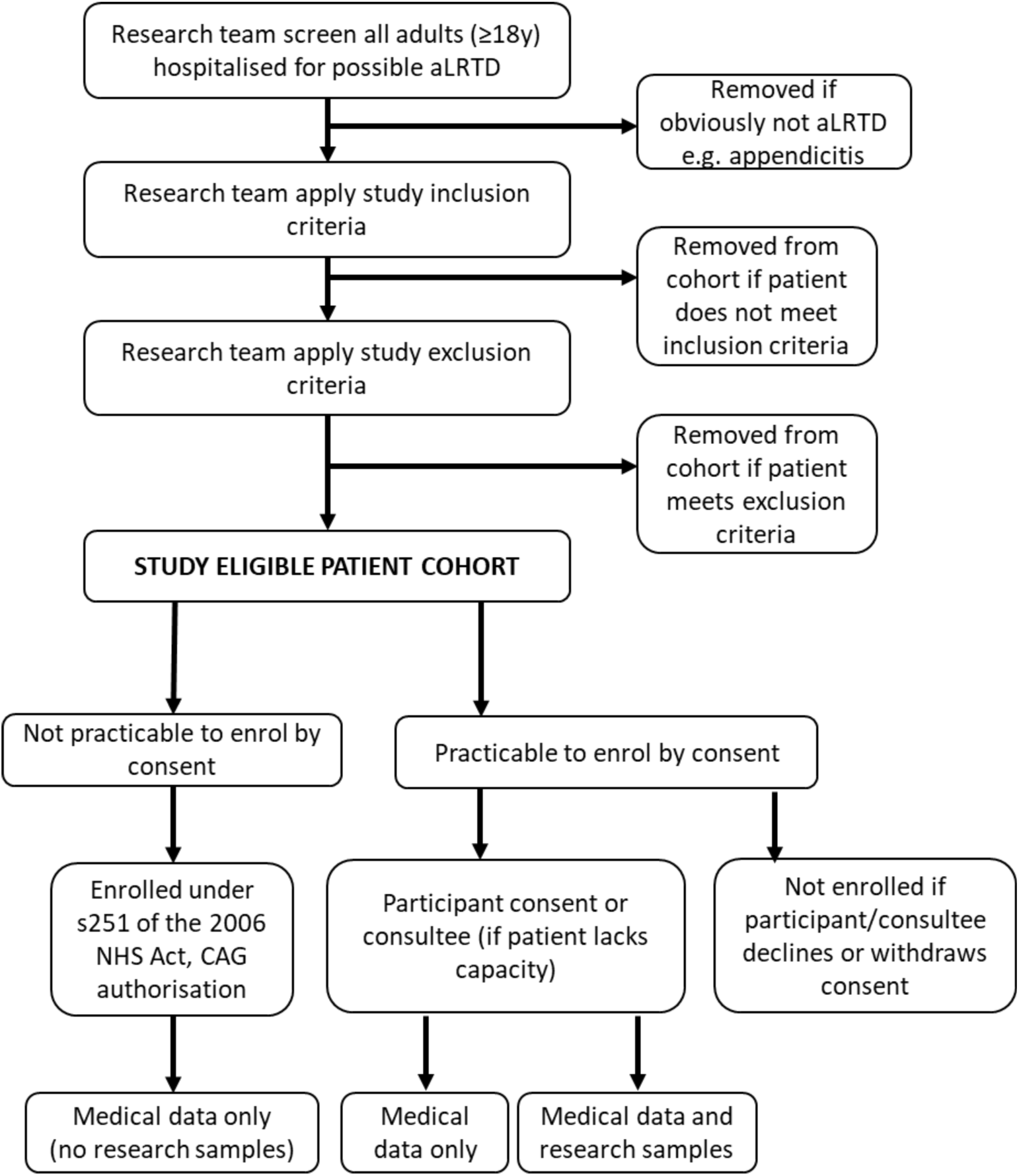
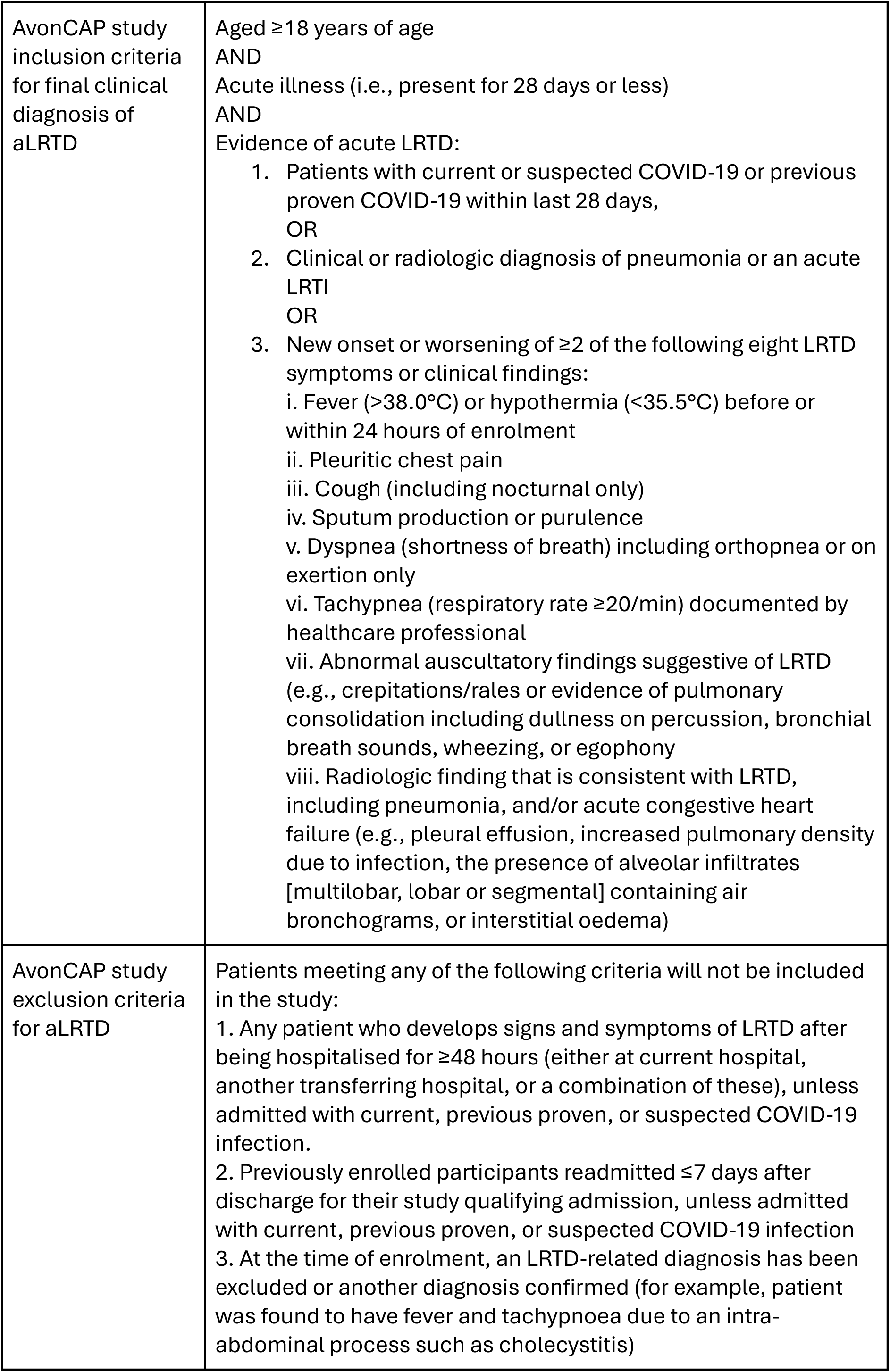
Enrolment procedures in The AvonCAP Study (ISRCTN 17354061) AvonCAP is a prospective observational cohort study, which undertakes comprehensive surveillance of aLRTD on all adults hospitalised in a defined geographical area (Bristol, UK). The study protocol is published and available at https://www.isrctn.com/ISRCTN17354061. The research team screens all adults (aged ≥18y) hospitalised at North Bristol NHS Trust and University Hospitals of Bristol and Weston NHS Trust for signs/symptoms of aLRTD, or a radiologically/clinically confirmed diagnosis of aLRTD. Patients whose symptoms are attributable to other conditions (e.g. pancreatitis, urosepsis) are not included, nor are patients who have hospital-acquired infection. Once the research team has determined that a patient is eligible for enrolment in the study (i.e. has aLRTD), they endeavour to approach patients wherever possible to ask for informed consent to enrol in the study. One of the following study processes may then occur: **1.** It is not practicable to enrol the participant by consent. The patient into a data collection study arm, using specific legal authorisation approved by the Confidentiality Advisory Group (CAG) under Section 251 of the 2006 NHS Act. Examples of this include: patients who are dead at the time of enrolment, patients who are discharged at the time of enrolment and who the study team cannot contact **2.** The patient has capacity and is enrolled by consent The research team obtain informed consent from the participant for enrolment in the AvonCAP study. To participate in the study, participants must give consent for the research team to use their medical data, but have the option to consent to research samples being taken (and thus may be enrolled but not provide research samples). **3.** The patient lacks capacity and is enrolled through a personal or nominated consultee If the patient lacks capacity at the time of enrolment, the research team obtains a declaration from a personal or professional consultee and if appropriate enrol the participant, in line with HRA guidelines. Enrolment includes the use of medical data, but research samples are optional (and thus participants may be enrolled but not provide research specimens). The research team monitors the participant during their hospitalisation to determine if they regain capacity. If this happens, the research team approaches the participant and obtains consent from the individual which supersedes the declaration. This may occur if patients have temporary delirium or are intubated on ITU and subsequently extubated. **4.** The patient or consultee declines consent, or it is withdrawn after enrolment The patient is not enrolled in the study if consent is declined. If they are withdrawing consent, they are withdrawn from the study and they are asked whether they wish for us to delete any obtained data and destroy research samples. The study then acts inline with the wishes of the participant. This process is outlined in the flow diagram below, and study inclusion/exclusion criteria are provided in a table for information.

**SUPPLEMENTARY DATA 2:**
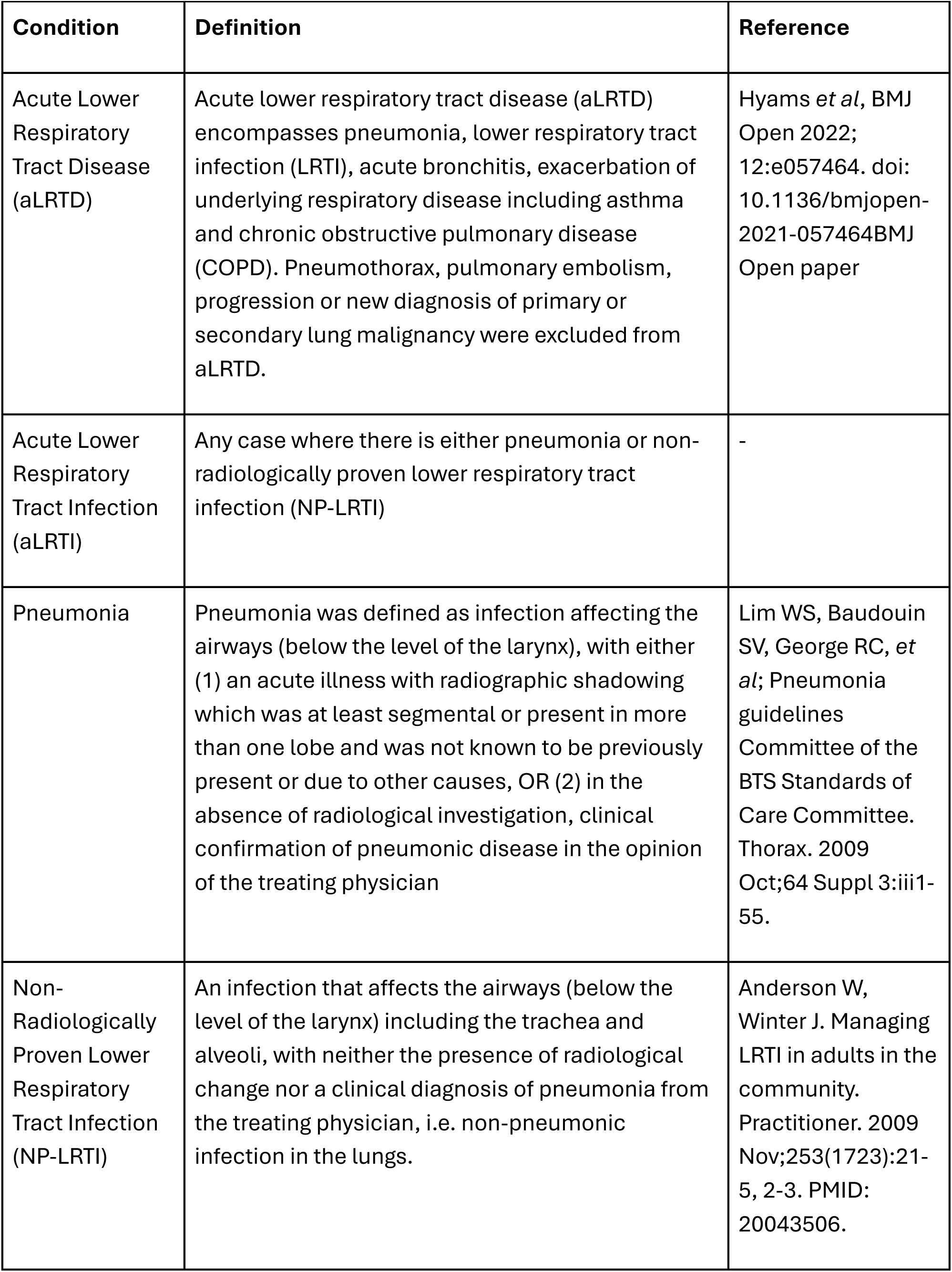
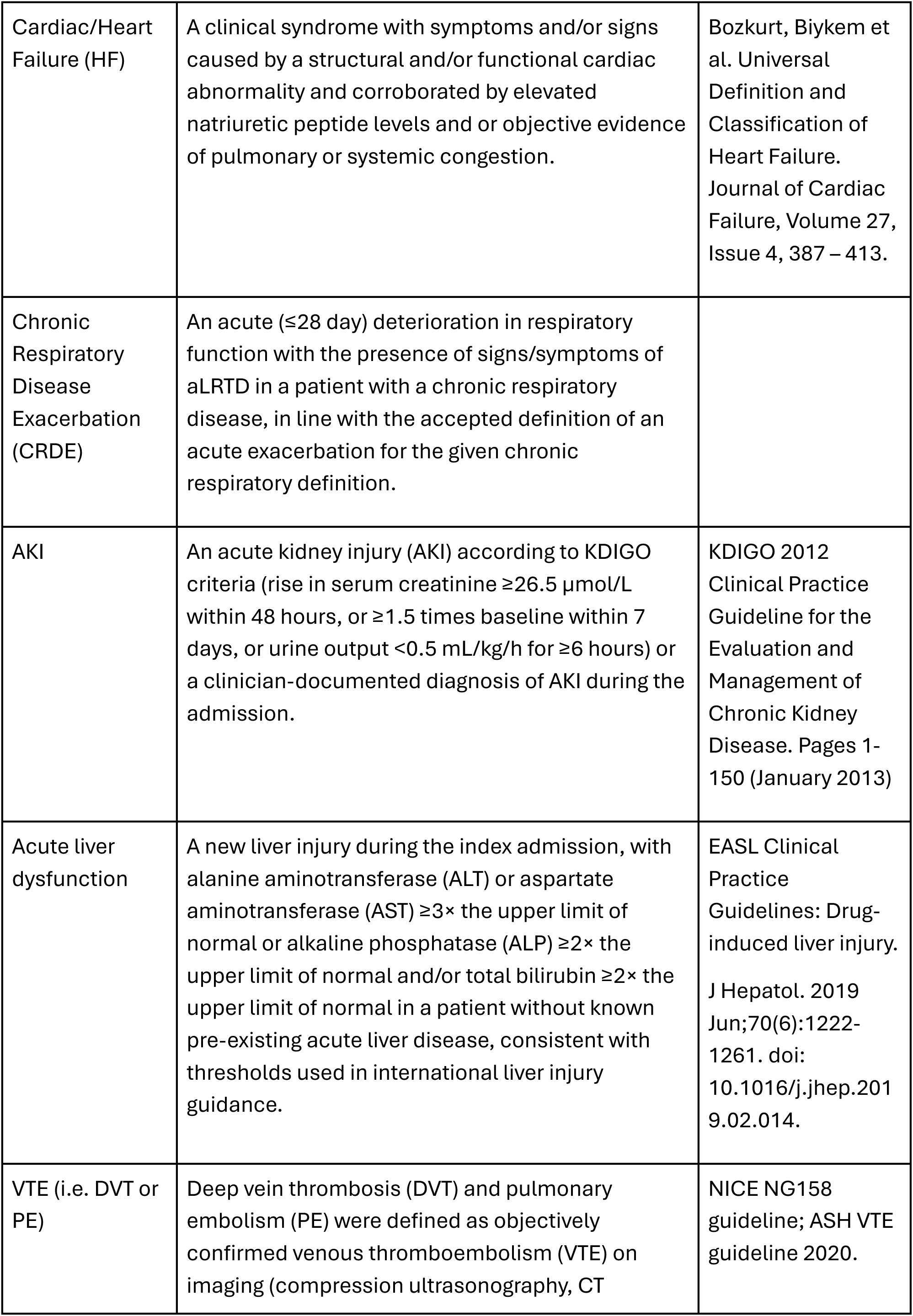
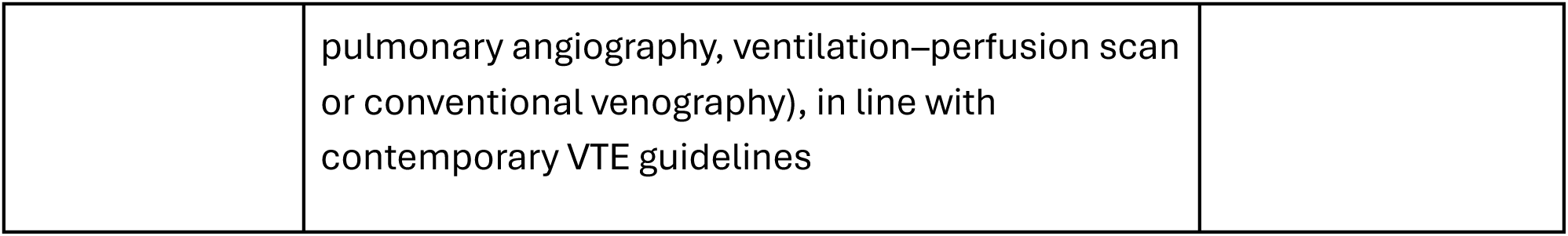
CASE DEFINTIONS.

**SUPPLEMENTARY DATA 3:**
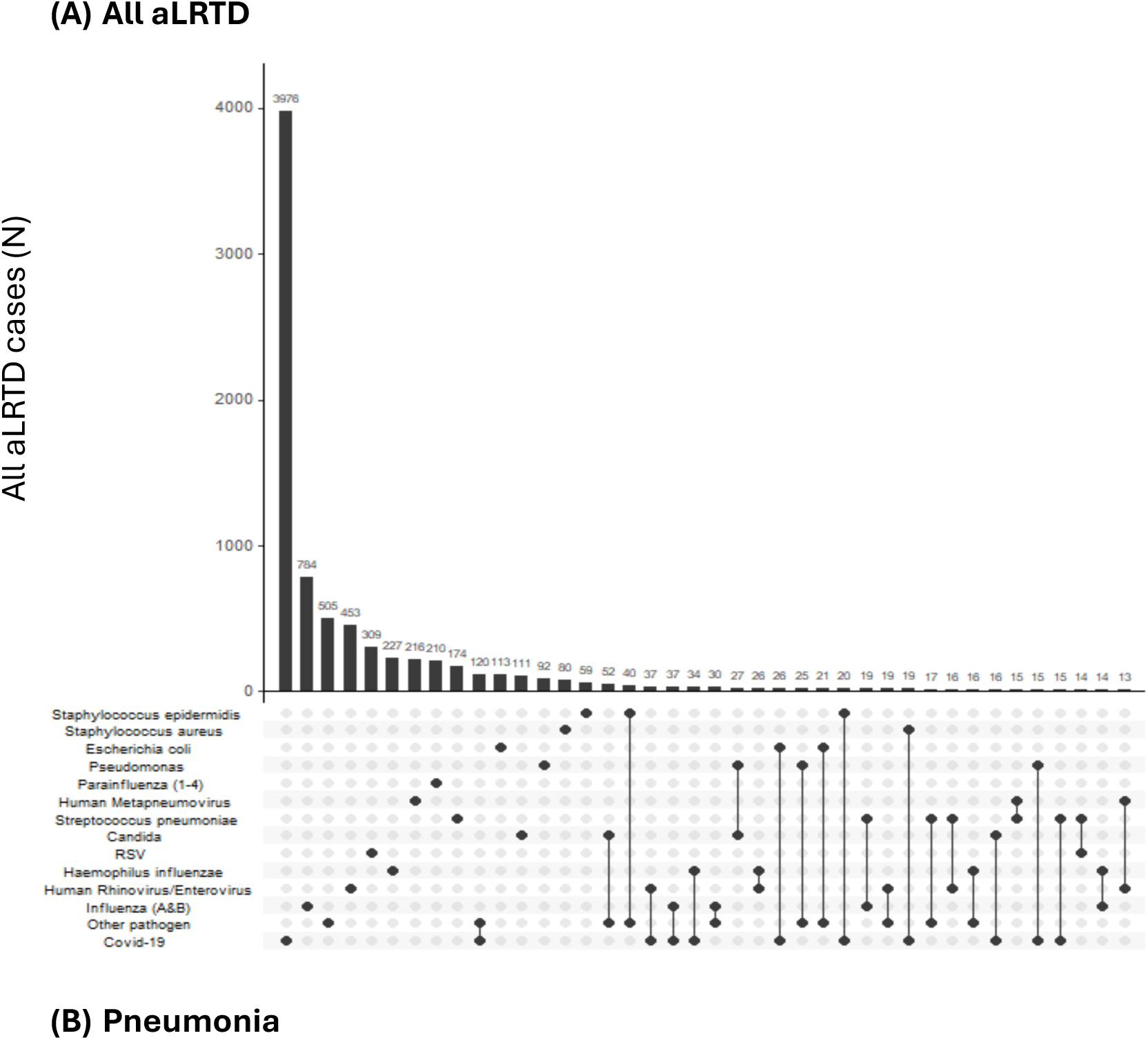
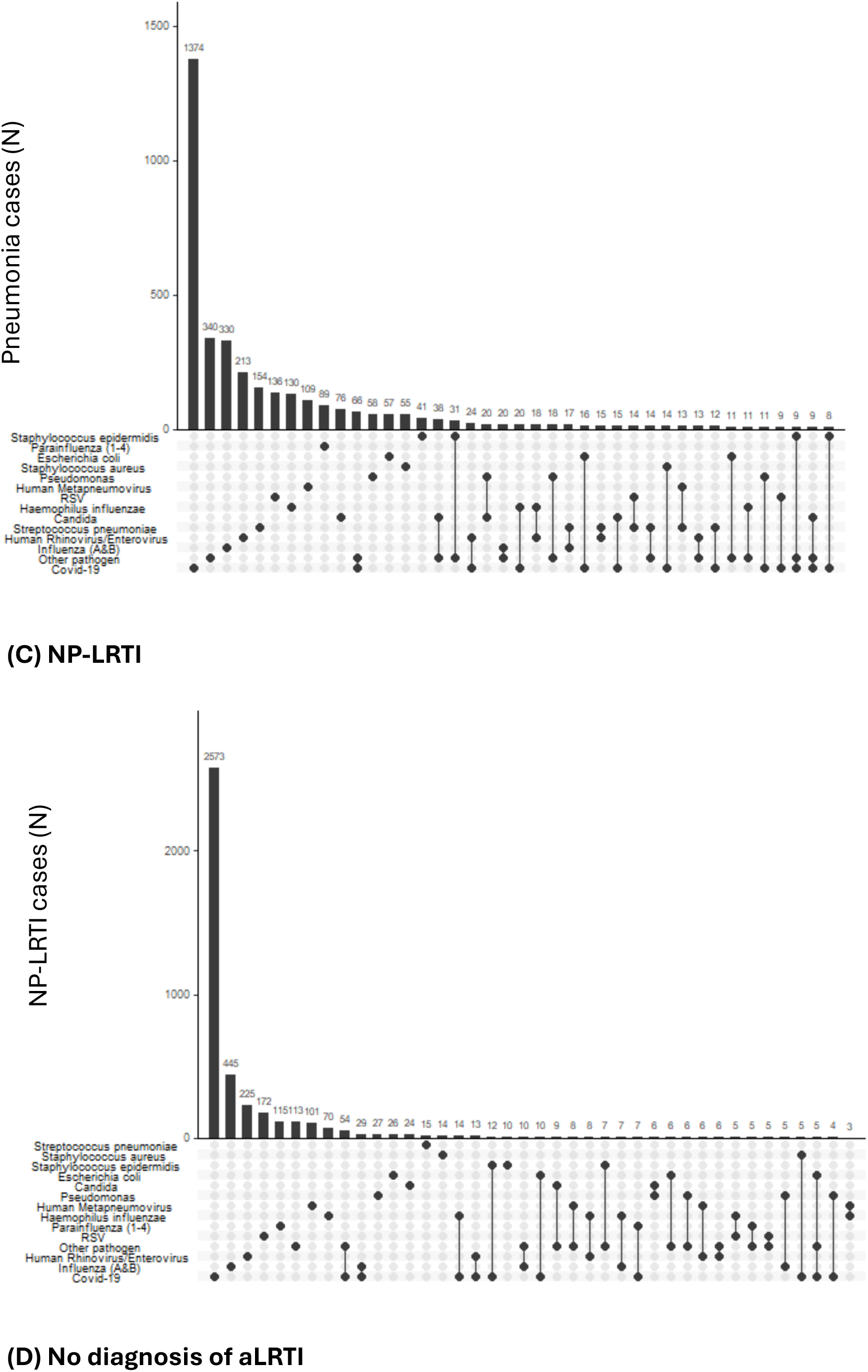
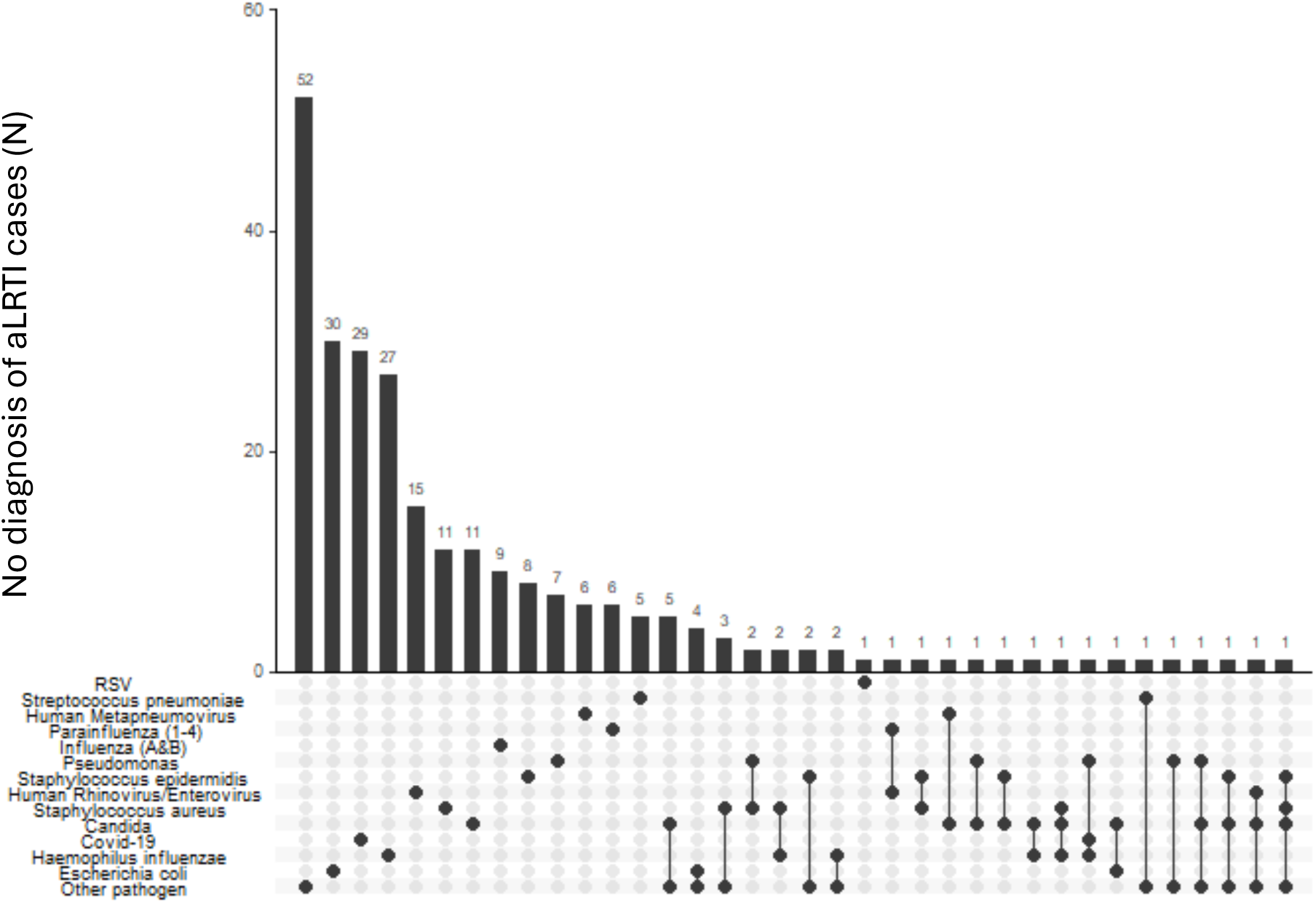
UpSet Plots of Respiratory Pathogen Co-Infections in adults hospitalised with aLRTD.

**SUPPLEMENTARY DATA 4:** Microbiological and virological testing among adult hospital aLRTD cases, stratified by pneumonia, non-pneumonic lower respiratory tract infection (NP-LRTI), and those with no aLRTI diagnosis. Values are numbers and column percentages of admissions in which each test type was performed, within each group. Testing includes both standard-of-care and research microbiological and virological samples. A single admission may have undergone multiple test types. ‘Tested – type not recorded’ refers to admissions with documented testing but no test type recorded. ‘Not tested’ refers to admissions with no microbiological or virological testing recorded.

| Test type | All aLRTD<br>N=21456 | Pneumonia<br>N=10239 | NP-LRTI<br>N=7742 | No aLRTI diagnosis<br>N=3475 |
| --- | --- | --- | --- | --- |
|  | N (%) | N (%) | N (%) | N (%) |
| Blood culture | 8147 (38.0%) | 5087 (49.7%) | 2420 (31.3%) | 640 (18.4%) |
| Sputum | 3540 (16.5%) | 2243 (21.9%) | 1028 (13.3%) | 269 (7.7%) |
| Urinary antigen - pneumococcus | 1938 (9.0%) | 1720 (16.8%) | 179 (2.3%) | 39 (1.1%) |
| Urinary antigen - legionella | 2073 (9.7%) | 1826 (17.8%) | 197 (2.5%) | 50 (1.4%) |
| Pleural fluid | 252 (1.2%) | 224 (2.2%) | 4 (0.1%) | 24 (0.7%) |
| Bronchoalveolar lavage | 52 (0.2%) | 48 (0.5%) |  | 4 (0.1%) |
| Tracheal Aspirate | 59 (0.3%) | 52 (0.5%) | 5 (0.1%) | 2 (0.1%) |
| Respiratory swab | 292 (1.4%) | 188 (1.8%) | 85 (1.1%) | 19 (0.5%) |
| PCR - COVID only | 2345 (10.9%) | 1193 (11.7%) | 891 (11.5%) | 261 (7.5%) |
| PCR Respiratory (Biofire) | 6977 (32.5%) | 3828 (37.4%) | 2451 (31.7%) | 698 (20.1%) |
| PCR - Respiratory panel | 2323 (10.8%) | 1204 (11.8%) | 935 (12.1%) | 184 (5.3%) |
| PCR Quad Panel | 4379 (20.4%) | 2251 (22.0%) | 1399 (18.1%) | 729 (21.0%) |
| Lateral Flow - COVID only | 224 (1.0%) | 97 (0.9%) | 103 (1.3%) | 24 (0.7%) |
| POCT Test (Abbott) | 11770 (54.9%) | 5775 (56.4%) | 4537 (58.6%) | 1458 (42.0%) |
| Tested - type not recorded | 6 (0.0%) | 1 (0.0%) | 2 (0.0%) | 3 (0.1%) |
| Not tested | 2304 (10.7%) | 735 (7.2%) | 623 (8.0%) | 946 (27.2%) |

